# Longitudinal Comorbidity Patterns Prior to Pulmonary Hypertension Diagnosis

**DOI:** 10.1101/2025.08.13.25333632

**Authors:** Mohsin Masood, Hong Pan, Juan Delgado-SanMartin, Siying Jane Ong, Xiaohe Zhang, Naailah Bhamji, Roger Thompson, Allan Lawrie, Dennis Wang

## Abstract

**BACKGROUND:** Pulmonary hypertension (PH) is a progressive and life-threatening condition that is often diagnosed late due to nonspecific symptoms shared with other cardiopulmonary diseases. The temporal relationship between comorbidities and PH diagnosis remains poorly understood, particularly regarding how disease order may influence diagnosis delays and prognosis.

**METHODS:** Pulmonary hypertension (PH) is a progressive and life-threatening condition that is often diagnosed late due to nonspecific symptoms shared with other cardiopulmonary diseases. The temporal relationship between comorbidities and PH diagnosis remains poorly understood, particularly regarding how disease order may influence diagnosis delays and prognosis.

**RESULTS:** The most common comorbidities among PH patients included hypertension (59.1%), heart failure (43.2%), and atrial fibrillation (31.8%). Pathway analysis revealed that PH diagnosis occurred 2.75 years earlier when diabetes preceded hypertension compared to the reverse order. In another pathway, advancing the position of hypercholesterolemia before hypertension and diabetes significantly shortened the progression time to PH across all three disease cohorts. Mortality risk varied by sequence type; a cardiometabolic trajectory involving diabetes, hypertension, and hypercholesterolemia was associated with a significantly higher mortality risk (HR = 1.97; *P* = 0.001). The relative order of metabolic conditions and comorbidity clusters was also associated with mortality.

**CONCLUSIONS:** Comorbidity order and sequence significantly affect the timeline and risk associated with PH diagnosis. Our findings demonstrate that earlier occurrence of certain metabolic conditions may determine disease progression speed and prognosis. These results support the utility of temporal disease trajectory modeling for risk stratification and early intervention of patients at risk for PH.

**Clinical Perspective:** *WHAT IS NEW?:*

- Using process mining, we identified how the order and sequence of comorbidities influence the speed of pulmonary hypertension (PH) diagnosis in a large real-world UK Biobank cohort.
- Patients who were diagnosed with type 2 diabetes before hypertension reached a PH diagnosis significantly faster (by 2.75 years) than those with reversed comorbidity order.
- Certain metabolic pathways were associated with significantly higher mortality risk (HR = 1.97), highlighting the prognostic importance of comorbidity sequences in PH.

*WHAT ARE THE CLINICAL IMPLICATIONS?:*

- The sequence of cardiometabolic comorbidities can impact diagnostic delays and survival outcomes in PH.
- This temporal patterning could support earlier diagnosis or risk stratification of patients suspected of having PH.
- Incorporating comorbidity sequence logic into electronic health record systems may help improve clinical decision support for timely PH diagnosis.

---

Pulmonary Hypertension (PH) is a chronic cardiovascular disease described by high blood pressure in the pulmonary arteries. PH affects the quality of life and reduces life expectancy ^1, 2^ and regularly develops in conjunction with heart failure and chronic lung conditions ^3, 4, 5^. The initial symptoms of PH including breathlessness, fatigue, and dizziness may lead healthcare professionals to misdiagnose patients as having other conditions such as asthma or heart failure (HF), resulting in significant diagnostic delays ^6, 7^. Right heart catheterization to measure pulmonary artery pressure stands as the best method to confirm PH diagnosis, but its invasive and specialized nature prevents it from serving as an initial screening procedure ^8^. Survival rates vary between different subgroups of PH ^9^, and previous studies have shown even mildly elevated mean pulmonary artery pressure of 19-24mm Hg are associated with an increased risk of death. Falling within this borderline range has redefined earlier diagnostic guidelines to emphasize the importance of early detection and management ^10, 11, 12^. Yet, patients suspected of having PH may need multiple visits to primary healthcare providers and specialists to undergo numerous medical tests, before a diagnosis is reached, particularly in cases of pulmonary arterial hypertension (PAH), where symptoms are often non-specific and diagnosis comes from by exclusion ^13^. A survey from the Pulmonary Hypertension Association UK (PHA-UK, 2023) revealed that one-third of PH patients waited over three years for diagnosis confirmation ^14^. To reduce the diagnostic delay, medical professionals should enhance awareness and screening of patients so that the most at risk of PH receive diagnostic tests earlier ^7, 8^.

Population-based cohort studies have faced persistent challenges in accurately identifying pulmonary hypertension (PH) patients due to the limited granularity of ICD-10 codes. These codes do not differentiate between the five clinical subtypes of PH (Groups 1–5), often resulting in misclassification or overly broad groupings that hinder subtype-specific research and clinical interpretation ^15, 16^. Late detection is also common, as PH frequently presents alongside cardiovascular and respiratory comorbidities. This highlights the need to evaluate whether comorbidity analysis can aid in early detection and risk stratification ^17^. Prior studies ^18, 19^ have demonstrated that specific conditions such as hypertension, atrial fibrillation, heart failure, and ischemic heart disease, as well as respiratory disorders like chronic obstructive pulmonary disease (COPD), interstitial lung disease (ILD), and pulmonary fibrosis commonly co-occur in PH patients. However, identifying which comorbidities most influence PH diagnosis and disease progression remains a significant challenge ^18, 19^. Importantly, the relative timing of these comorbidities relative to diagnosis and each other may offer deeper insights into disease risk. These may also reveal opportunities for earlier intervention and tailored treatment strategies ^20, 21, 22^. Several studies have emphasized the importance of understanding the temporal sequence and clustering of comorbidities. For instance, findings from the COMPERA registry showed increasing comorbidity rates over time in PH patients, underscoring the need for adaptive treatment approaches ^23^. A 15-year longitudinal study by Santiago-Vacas et al. ^24^ tracked PH progression alongside right ventricular dysfunction in heart failure patients, providing valuable insights into long-term prognosis. Yet, these studies did not examine PH comorbidities longitudinally.

In this study, we explored the sequence and timing of comorbid conditions preceding a PH diagnosis using a data driven technique in business analytics, process mining, that has been successfully applied in different disease contexts including COVID-19 mortality prediction, stroke rehabilitation, heart failure management, and dementia care ^25, 26, 27, 28, 29^. We compared the comorbidity sequences both before and after PH diagnosis with matched cohorts of heart and lung diseases. To assess the molecular underpinnings of PH comorbidities, we incorporated publicly available GWAS summary statistics and appliedy linkage disequilibrium score regression (LDSC) to estimate genetic correlations between PH and its major comorbidities. Finally, we estimated the increased risk of mortality associated with the different comorbidity sequences.

## METHODS

This study was conducted using the UK Biobank data set under approved access (Application Number: 85457). Ethical approval was obtained from the Northwest Multi-center Research Ethics Committee (REC reference: 11/NW/0382), and all participants provided informed consent at recruitment. The data cleaning, preprocessing, and process mining stages of the analysis were implemented in Python (<u>v 3.9.12</u>) and R (<u>v</u> <u>4.3.2</u>). The complete set of experiments, including the full analysis codebase for data processing, comorbidity trajectory mining, and figure generation, is permanently archived and freely accessible at: https://doi.org/10.5281/zenodo.15747766 and <u>GitHub</u>.

### Identifying a PH Cohort

From the 502,499 UK Biobank participants with electronic health records under our application ID 85457, we extracted the ICD-10 codes dataset, containing diagnostic information for 440,017 participants (covering the period from 1992 to 2021), and the all-cause mortality dataset, which included death records for 37,897 participants and their detailed causes of death for 37,731 (99.5%) of them. After removing duplicates, 3 participants were excluded due to missing key information (e.g., incomplete diagnostic records or inconsistent ICD-10 codes). This resulted in a final data set comprising 440,014 participants. From this, we identified a cohort of 2,727 PH patients. We subdivided the population into three primary classes: Primary PH (I27.0, n=776), Other Secondary PH (I27.2, n=1,949), and Secondary PH (I27.9, n=395). This PH cohort of 2,727 included 1,451 males (53.21%) and 1,276 females (46.79%), differing slightly from the broader UK Biobank population, where males comprised 45.17% and females 54.83%. A full breakdown of the cohort characteristics is provided in Supplementary Material (see Table I). The ICD-10-based classifications for PH can be contextualized within the broader PH group classification (Groups 1-5) defined by the 6th World Symposium on Pulmonary Hypertension (WSPH). Group 1 (PAH) aligns with the ICD-10 category of Primary PH (I27.0), while Groups 2-5 correspond to various forms of secondary PH, including PH due to left heart disease (Group 2), lung diseases or hypoxia (Group 3), chronic thromboembolic PH (Group 4), and multifactorial mechanisms (Group 5) ^1, 10^.

### Identifying comorbidities in the PH cohort, and in matched COPD and heart failure cohorts

We further extracted the most frequently co-occurring ICD-10 diagnoses to identify comorbidities within this population before categorizing them at the subchapter level. In the PH patient group consisting of 2,727 participants, essential hypertension was also present in 1,611 (59.1%), atrial fibrillation in 868 (31.8%), COPD in 1,822 (66.8%), heart failure in 1,179 (43.2%), hypercholesterolemia in 830 (30.4%), and diabetes mellitus in 579 (21.2%) patients. Since COPD and HF are thought to have related pathology to PH, we examined their comorbidities as comparable controls to the comorbidities observed for PH. We extracted a COPD-matched cohort and a heart failure (HF)-matched cohort, each consisting of 2,441 participants who had never been diagnosed with PH. A detailed description of the control cohort selection and matching methodology is provided in the Supplementary Material (Section 1.5).

### Assessing Genetic Overlap Between PH and Comorbidities Using GWAS Data

We used linkage disequilibrium score regression (LDSC v1.0.1) to evaluate whether PH and its comorbidities share common genetic risk factors by estimating SNP-based heritability and pairwise genetic correlations. This research extracted common genetic variants called SNPs from publicly available GWAS summary statistics while concentrating on European-ancestry groups and high-quality HapMap3 SNPs. We excluded comorbid traits that showed very low heritability levels below 1% and those with non-significant correlation results exceeding a p-value of 0.05. Supplementary Methods (Section 4) contain detailed descriptions of dataset sources, quality control filters, and LDSC parameters used in our methodology.

### Process mining of comorbidity progression over time

We extracted dates of diagnostic events from hospital inpatient records and primary care data in the UK Biobank which links each patient’s ICD-10 diagnosis to the date they were diagnosed or hospitalized. The initial occurrence date was used to timestamp all ICD-10-coded diagnoses for each participant. We further identified the PH diagnosis date by selecting the first recorded date among all ICD-10 codes related to PH which include I27.0, I27.2, and I27.9. Comorbidities are sorted into chronological order based on the PH diagnosis date, then separated into pre-PH (diagnosed before PH) and post-PH (diagnosed after PH) categories. We utilized process mining techniques in the <u>bupaR</u> package in R to create sequences of comorbidities that describe the order and time between diagnoses. The ordered sequences referred to as pathways were visualized as event logs to illustrate comorbidity progression routes that lead to PH, COPD, and HF.

### Estimating mortality risk for different comorbidity pathways

To evaluate mortality risk associated with each identified comorbidity pathway, we used linked mortality data from the UK Biobank. We performed time-to-event analysis using Cox proportional hazards models, estimating hazard ratios (HR) to quantify the risk of death associated with specific comorbidity trajectories. Age (at PH diagnosis), and sex as covariates were included as adjustment variables where applicable (details in Supplementary Material section 1.4, 1.6).

## RESULTS

### Pulmonary hypertension has a similar profile of pre-diagnosis comorbidities as heart failure

There were 2,727 individuals in UK Biobank who received a PH diagnosis from 1995 to 2021 as described by their ICD-10 codes. We classified comorbid conditions into three types based on their diagnosis timing relative to the assignment of a PH diagnosis code (I27.0, I27.2, I27.9): pre-PH comorbidities (diagnosed only before PH diagnosis), post-PH comorbidities (diagnosed only after PH diagnosis), and common PH comorbidities (diagnosed both before and after PH diagnosis). To eliminate duplication, we included only the first recorded instance of each comorbidity. The distribution of comorbidities was similar to existing comorbidities found at specialized pulmonary hypertension centers (Fig.1) ^30^. For the COPD matched control cohort, the most prevalent comorbidities included chronic ischemic heart disease with 1,377 cases representing 56.4%, hypertension with 1,138 cases equating to 46.6%, along with hypercholesterolemia which occurred in 543 cases or 22.2%. A total of 1,746 individuals in the HF matched cohort suffered from hypertension (71.5%), followed by hypercholesterolemia in 1,042 individuals (42.7%) and atrial fibrillation in 803 individuals (32.9%).

**Figure 1.**
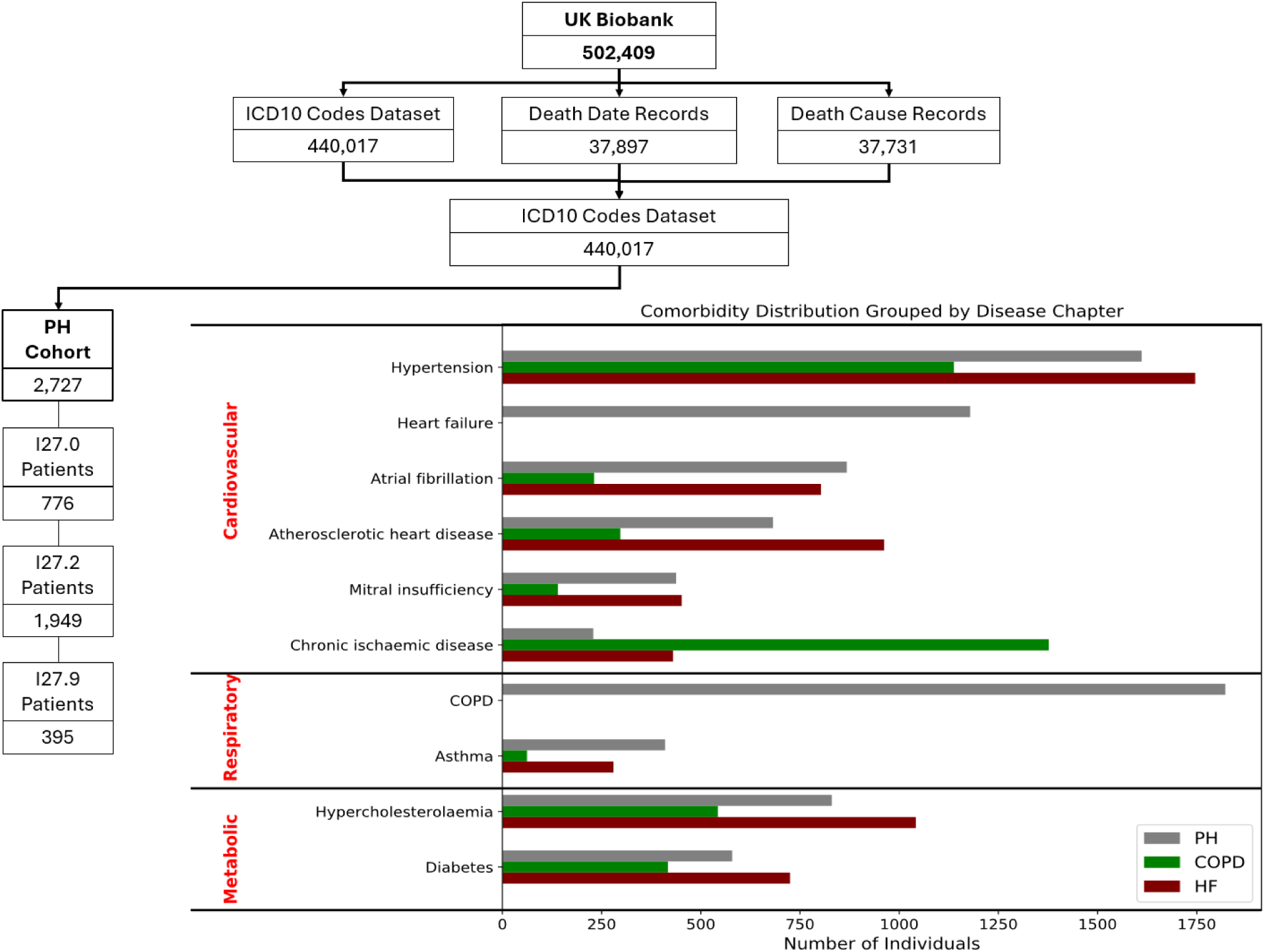
Study Flow and Derivation of PH Cohort and Comorbidity Profiles in PH, COPD, and HF Matched Control Groups: This figure outlines the preprocessing pipeline from the UK Biobank (n = 502,409) to the identification of 2,727 PH patients, stratified by ICD-10 codes into I27.0, I27.2, and I27.9 classes. The bar plot visualizes the distribution of key comorbidities among the PH cohort, COPD-matched controls, and HF-matched controls, categorized into three ICD-10 disease chapters: Cardiovascular, Respiratory, and Metabolic. Horizontal black lines delineate the chapter groupings, with comorbidities arranged in a manually defined order within each chapter.

Fig. 2 illustrates the distribution of comorbidities diagnosed before and after the index conditions (PH, HF, and COPD). All comorbidities are diagnosed before their respective index conditions, which suggests these conditions act as contributing factors rather than effects of those diseases. The most common comorbidity found before diagnosis was essential hypertension with a prevalence of 58.1% in PH patients, 38.5% in COPD patients and 69.7% in HF patients. Atrial fibrillation and flutter showed high prevalence in PH patients (31.2%) and HF patients (31.3%) while remaining significantly less common in COPD patients (6.5%, *χ²* (2) = 568.07, *p* < 0.0001). Asthma was most prevalent in PH patients (15.8%), compared to both COPD (1.6%) and HF (9.8%). PH patients exhibited a pattern of cardiovascular comorbidities, including hypertension, atrial fibrillation, atherosclerosis, and hypercholesterolemia, that was more similar to that of HF patients than to COPD patients. Unlike COPD patients, who showed the highest prevalence of chronic ischemic heart disease (56.4%), PH patients had a lower prevalence (14.9%). The pre-diagnosis timing of comorbidities in PH patients followed a pattern that was more comparable to HF patients than to COPD patients.

**Figure 2.**
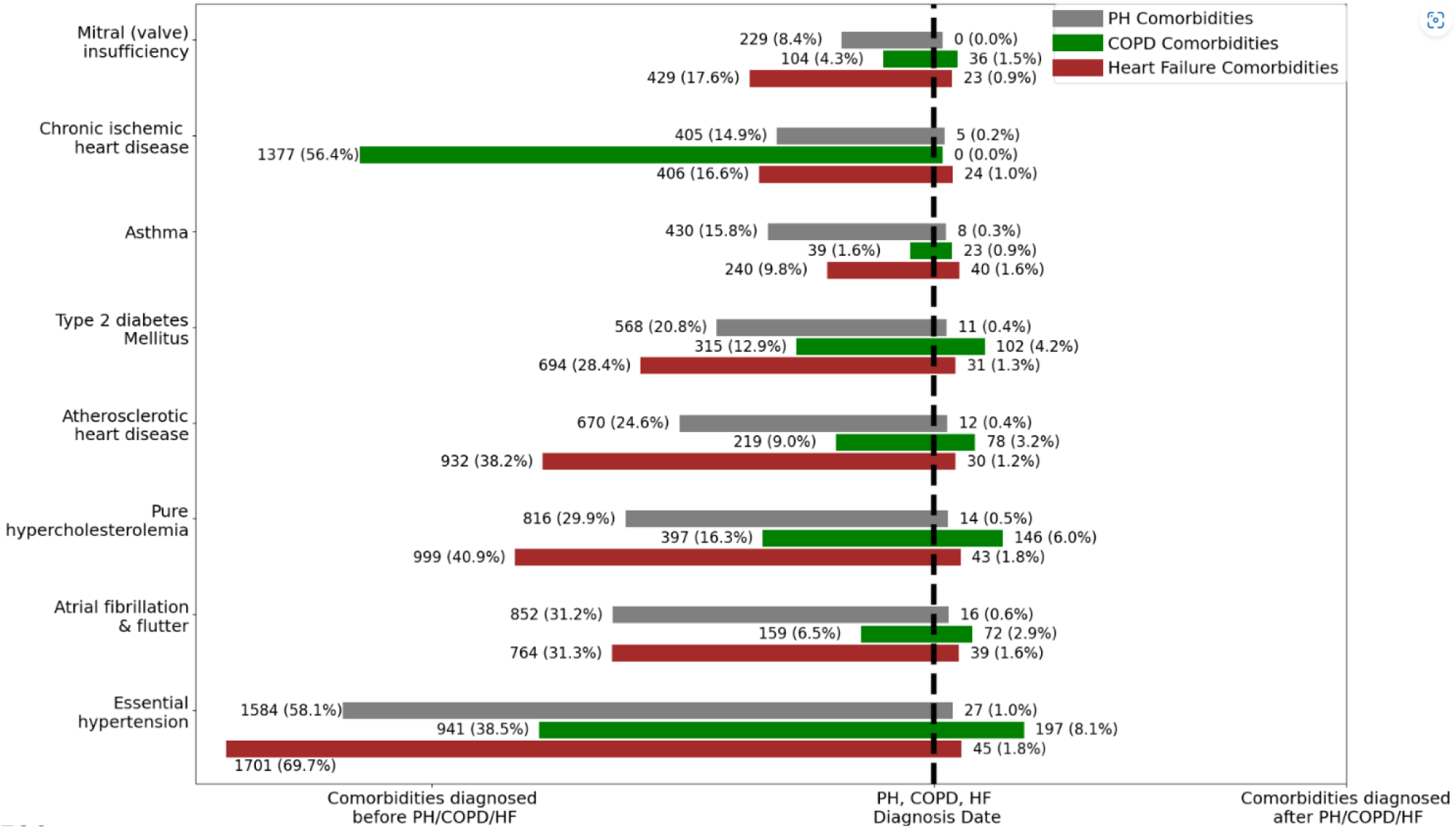
Timing and frequency of comorbidities in PH, and matched cohorts of COPD and HF: The forest plot depicts the frequency of various comorbidities occurring before, after, and on the same date with a diagnosis of PH. Each bar is labeled with the number of patients and percentage with the comorbidity in the respective cohort.

### Strong genetic overlap between PH and cardiovascular comorbidities

We observed the strongest genetic correlation between PH and heart failure, with an estimated r_g_=0.65 (SE = 0.18, P = 3.08 × 10^−4^). Notable correlations were also found with hypertension (r_g_ =0.62, SE=0.14, P=1.82× 10⁻^5^), hypercholesterolemia (r_g_ =0.52, SE = 0.14, P = 3.26 × 10^-4^), atrial fibrillation (r_g_ =0.64, SE = 0.17, P = 1.10 × 10⁻⁴), and coronary atherosclerosis (r_g_ =0.42, SE = 0.11, P = 1.38 × 10⁻⁴). A smaller but statistically significant correlation was identified between PH and type 2 diabetes (r_g_ =0.26, SE = 0.08, P = 1.92 × 10⁻³). In contrast, no significant genetic correlation was observed between PH and either COPD or asthma in the GWAS datasets analyzed.

### Process mining reveals comorbidity pathways to pulmonary hypertension diagnosis

Since the diagnosis of PH may not only be influenced by the presence of comorbidities but also by the order they arise, we used process mining to examine the comorbidities as a pathway leading to PH. Fig. 3 highlights the longitudinal progression of comorbidities in relation to PH, revealing key temporal sequences between conditions both before and after PH diagnosis. Hypertension is the most prevalent pre-PH comorbidity, with a direct pathway to PH observed in 565 patients over a median of 3.90 years. Two primary pathways were identified: (1) *hypertension → atrial fibrillation →* PH, where 235 patients developed atrial fibrillation 2.30 median years after hypertension onset. Atrial fibrillation persisted for 4.31 median years in 294 patients before PH diagnosis. (2) *hypertension → atherosclerotic heart disease →* PH, where 228 patients developed atherosclerosis after 2.66 years from hypertension onset, with a subsequent progression to PH occurring over 1.94 years in 297 patients.

**Figure 3.**
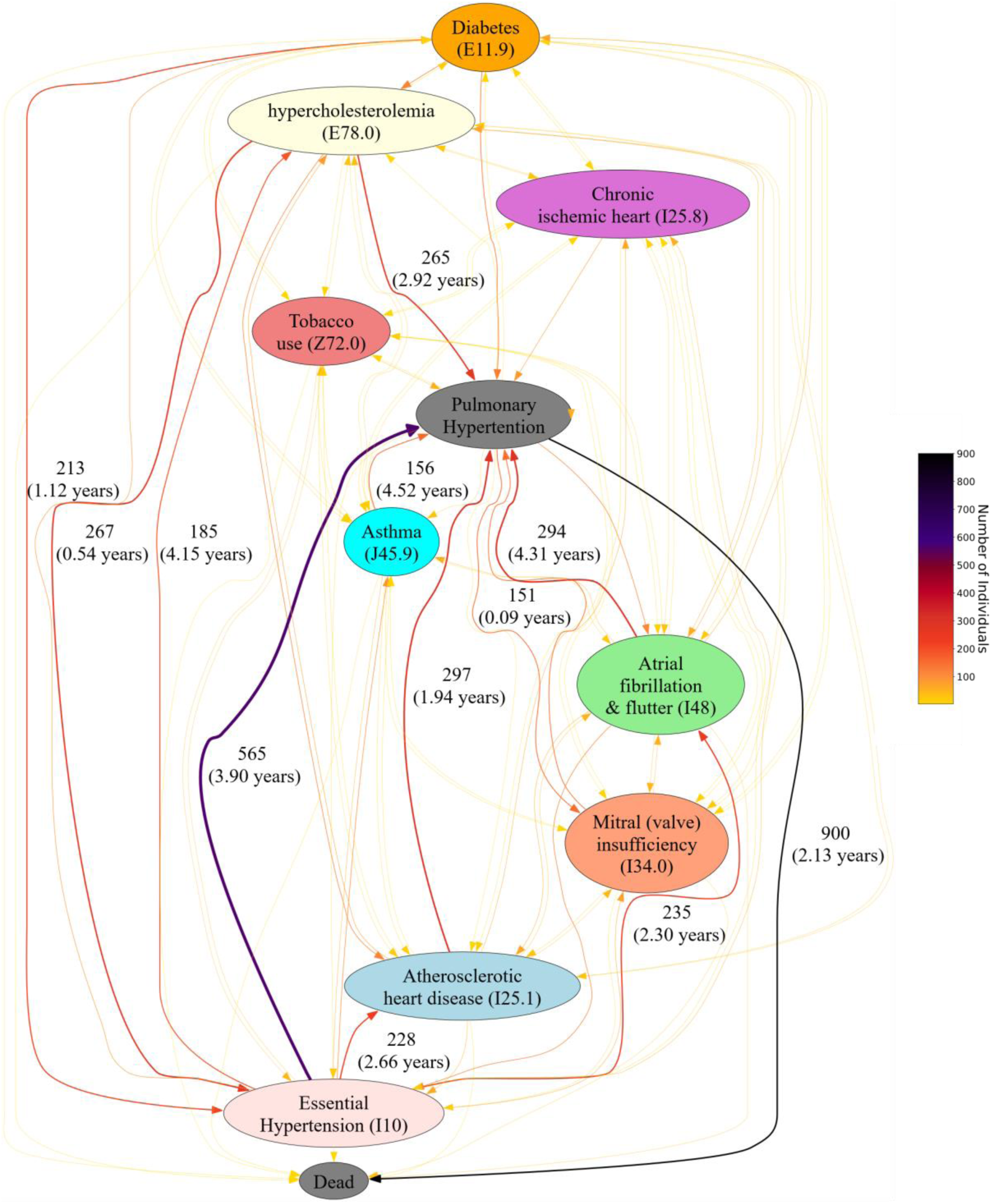
Process mining of longitudinal comorbidity pathways in PH cases: This figure illustrates the progression of comorbidities before and after PH diagnosis. Each node represents a condition, with connecting lines and arrows indicating transitions over time. The direction of the arrows reflects the sequence of progression, while the numbers adjacent to the lines represent the number of patients and the duration (in years - median) between stages. Line color intensity corresponds to patient frequency, emphasizing more prevalent pathways.

Respiratory and metabolic conditions such as asthma and hypercholesterolemia were frequently diagnosed prior to PH, with asthma persisting for 4.52 years in 156 patients before PH diagnosis and hypercholesterolemia present for 2.92 years in 265 patients. Following PH diagnosis, mitral valve insufficiency was frequently identified, with 151 patients receiving the diagnosis within a median of 0.09 years. This temporal proximity may reflect concurrent detection during diagnostic investigations such as echocardiography. Mortality was high, with 900 patients (out of 2,727 PH individuals) passing away within 2.13 median years after PH diagnosis.

### Temporal sequences of cardiometabolic comorbidities identify accelerated pathways to PH and reveal order-specific risks

To investigate whether the order of cardiometabolic comorbidities affects time to PH and other cardiopulmonary conditions, we examined matched sequences with the same conditions but different temporal arrangements (Fig. 4), prioritizing pathways that (i) show ≥2–3 year differences in median time to diagnosis and (ii) exhibit consistent trends across PH, COPD, and HF cohorts. A striking example is the pair of *diabetes → hypertension →* PH/COPD/HF versus its reverse *hypertension → diabetes →* PH/COPD/HF. In the PH cohort, the first pathway leads to PH in 5 years 11 months, whereas the reversed order results in PH in 8 years 8 months. This directionality effect is even more pronounced in the COPD cohort (3 months vs. 9 years 7 months) and HF (5 years 5 months vs. 10 years 3 months), indicating that the early onset of diabetes followed by hypertension significantly accelerates disease onset. Another illustrative pair is *diabetes → hypertension → hypercholesterolemia →* PH/COPD/HF versus *diabetes → hypercholesterolemia → hypertension →* PH/COPD/HF. Although both pathways involve the same set of metabolic and cardiovascular comorbidities, the ordering resulted in different progression speeds. In the PH cohort, the former progressed in 9 years and 2 months, while the latter took only 6 years and 9 months. Similar differences were observed in COPD (10 years 8 months vs. 5 months) and HF (8 years 9 months vs. 10 years 4 months) cohorts, emphasizing that the timing and positioning of *hyperlipidemia* in the trajectory alters disease dynamics. The shortest pathway identified was *hypertension → atherosclerotic heart disease →* PH/COPD/HF, leading to PH in just 4 years 11 months, compared to its reverse *atherosclerotic heart disease → hypertension →* PH/COPD/HF, which required 5 years 11 months. In the COPD cohort, this pathway also progressed rapidly (2 years 2 months vs. 1 year 5 months), whereas in HF, the reverse sequence took markedly longer (4 years 4 months vs. 10 years 1 month).

**Figure 4.**
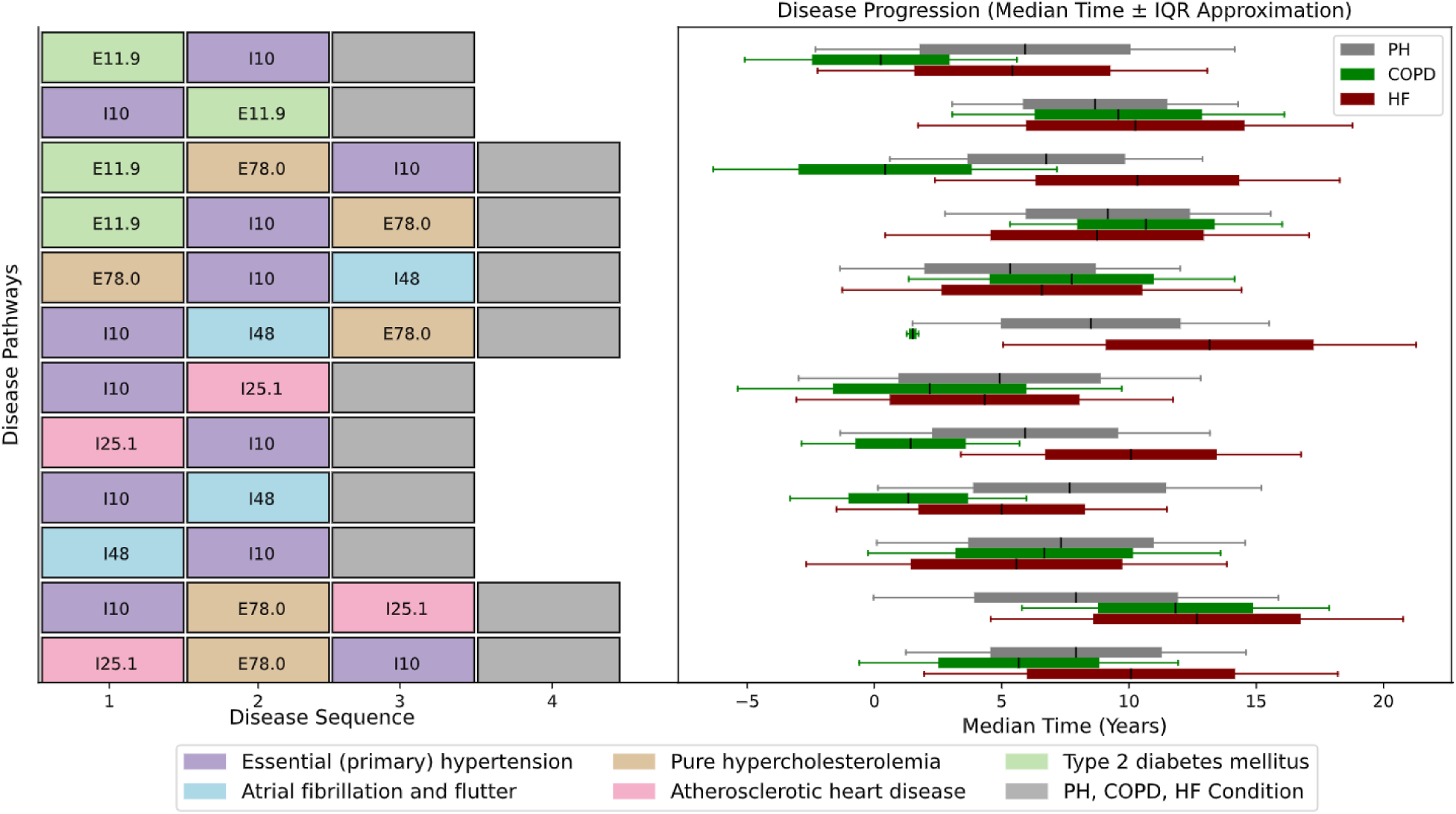
Comorbidity pathways and median time leading to diagnosis of PH, COPD, and HF cohorts: The cohorts had similar demographics and did not contain each other’s condition as comorbidity. The left panel visualizes comorbidity pathways preceding the diagnoses of pulmonary hypertension (PH), chronic obstructive pulmonary disease (COPD), and heart failure (HF), with each node representing a sequential ICD-coded condition. Pathways are grouped based on their initial comorbidity. The right panel presents horizontal boxplots showing the median time (in years ± IQR approximation) from the first condition in each pathway to diagnosis across PH, COPD, and HF cohorts, enabling comparative analysis of disease progression trajectories.

### Differences in PH mortality due to pre-diagnosis comorbidities

Given the different timelines of comorbidity pathways leading up to diagnoses, we hypothesized that this would also lead to different rates of disease progression for PH, COPD and HF. We assessed overall survival in patients grouped by their comorbidity pathways leading to PH and COPD (Fig. 5). For each pathway, Cox proportional hazards models were used to estimate mortality risk, comparing patients following that pathway to all other patients within the same disease group. The resulting hazard ratios also allowed us to compare which pathways were associated with higher mortality risk in PH versus COPD populations. With hazard ratios of 1.88 (p=0.001) for PH and 1.99 (p=0.010) for COPD, the *diabetes* → *hypertension* → *hypercholesterolemia* → PH/COPD pathway had similar mortality risk for both conditions. *hypercholesterolemia* → *hypertension* → *atrial fibrillation* → COPD is a pathway that increases the risk of death in people with COPD (HR= 2.51, p=0.001). In contrast, this same sequence does not show elevated risk in the PH cohort. The route with the highest mortality risk among all of them is *hypertension → diabetes → hypercholesterolemia →* COPD (HR= 3.67, p=0.001). A better prognosis was observed with the same sequence of metabolic comorbidities leading to PH also shows increased mortality (HR = 1.67, *p* = 0.001). A considerable mortality risk is shown by the pathway *atherosclerotic heart disease → hypertension →* COPD (HR= 2.27, p=0.010), while the same pathway leading to PH does not show the same mortality risk (HR=1. 39, p=0.16).

**Figure 5.**
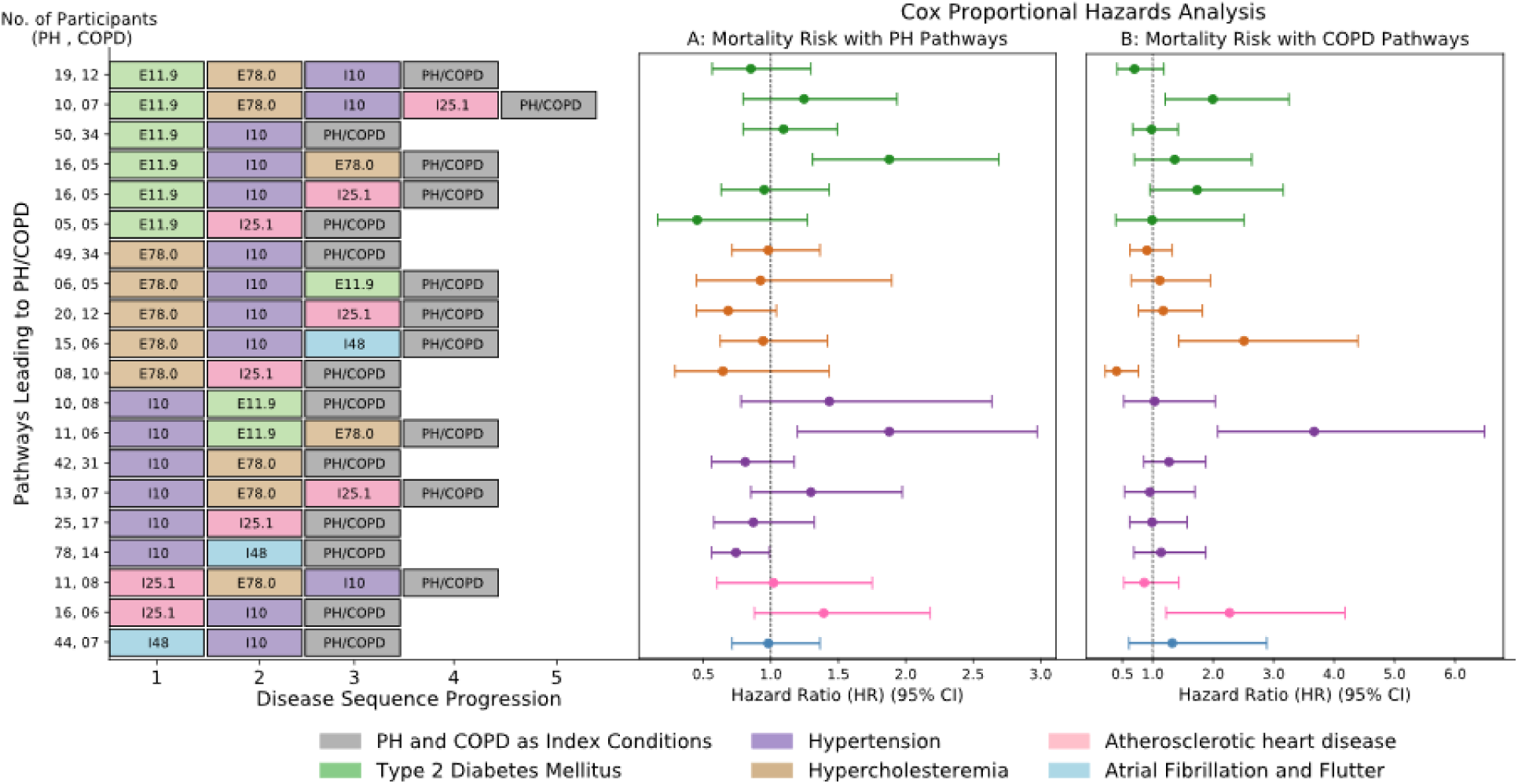
Pathways Leading to PH and COPD with Mortality Risk Assessment Using Cox Proportional Hazards Analysis: The left panel illustrates the (Disease Sequential order) pathways, with nodes representing comorbid conditions indicating sequential progression. The colored bars represent each pathway according to the first occurred condition, with the number of participants (PH, COPD) listed for each pathway at y-axis. The right panels display hazard ratios (HR) with 95% confidence intervals for mortality risk associated with PH pathways (A) and COPD pathways (B).

This same comparison was performed to evaluate the mortality risks associated with various comorbidity pathways leading to PH and HF (Fig. 6). The pathway *diabetes → hypertension → hypercholesterolemia →* PH presents the highest mortality risk in PH (HR= 1.97, p=0.001). The same sequence of comorbidities also had a poor prognosis in HF patients with an HR of 1.34 and p-value at 0.010. There is a considerable mortality risk for HF patients with a prior comorbidity pathway of *hypercholesterolemia → hypertension → atrial fibrillation → HF* (HR= 1.60, p=0.005). In contrast, the pathway *hypertension → diabetes → hypercholesterolemia →* PH leads to the highest mortality risk for PH (HR= 2.05, p=0.001). These findings emphasize distinct high-risk multimorbidity sequences across disease cohorts. Notably, no single comorbidity pathway was found to be significantly associated with high mortality risk in both PH and HF cohorts.

**Figure 6.**
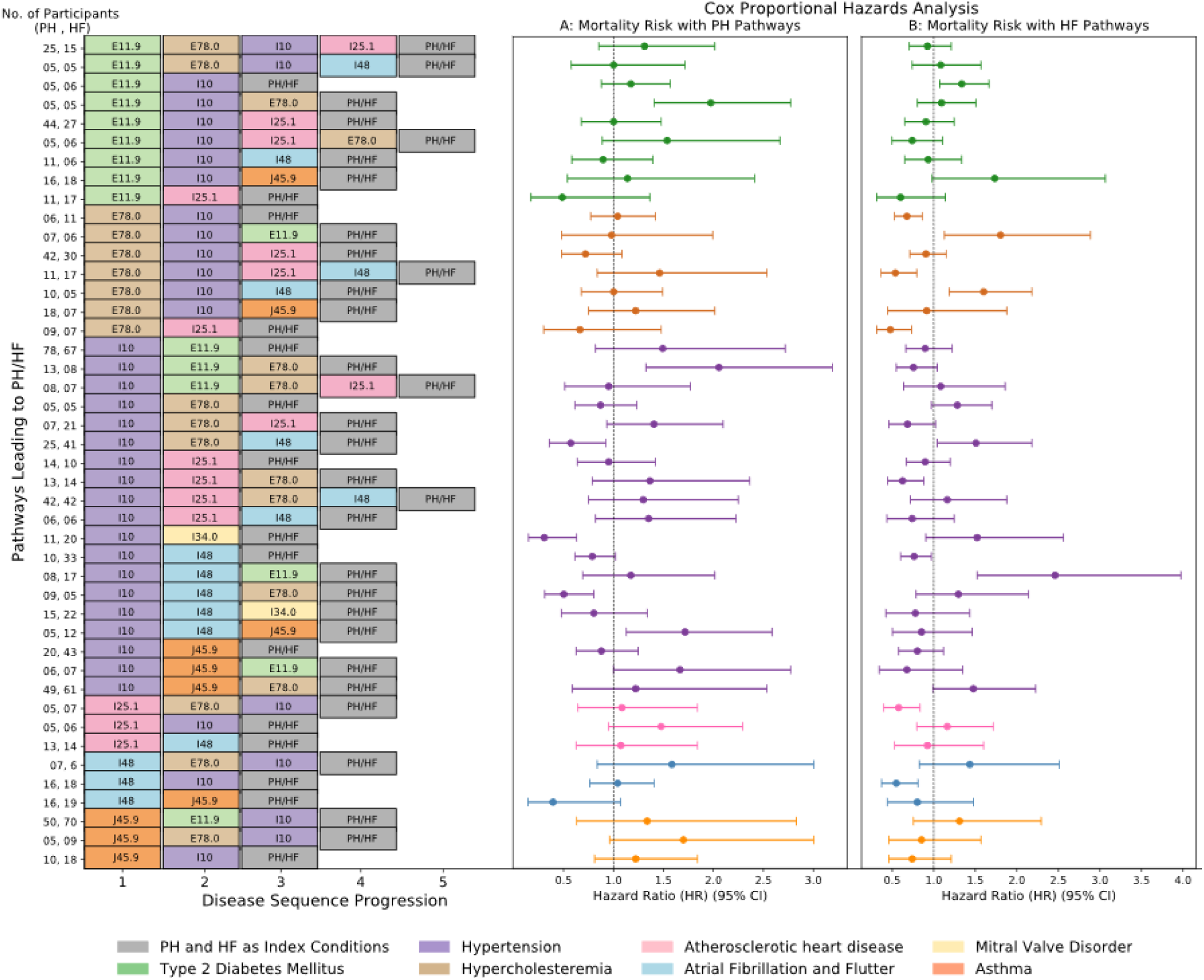
Pathways Leading to PH and HF with Mortality Risk Assessment Using Cox Proportional Hazards Analysis: The left panel illustrates the sequential order of diagnosis pathways, with nodes representing comorbid conditions indicating sequential progression. The colored bars represent each pathway according to the first occurred condition, with the number of participants (PH, HF) listed for each pathway at y-axis. The right panels display hazard ratios (HR) with 95% confidence intervals for mortality risk associated with PH pathways (first panel) and HF pathways (second panel).

## DISCUSSION

This study provides insights into the temporal progression and mortality risks of comorbidities in PH, HF, and COPD, leveraging UK Biobank data. The analysis identified clinically relevant temporal sequences of comorbidities through a dynamic analytical approach that surpasses traditional cross-sectional studies ^1^, ^29^. Hypertension affected 59.1% of patients, with atrial fibrillation (31.8%) and heart failure (43.2%) also highly prevalent. The distribution of the most common comorbidities across PH patients was similar to what was observed in a PH-specific disease cohort^30^. The similarity between the datasets confirms the strength and broad applicability of our comorbidity assessment in PH patients despite variations in population attributes and data origins. These findings reveal that cardiometabolic multimorbidity remains a significant problem in PH patients, especially in elderly and high-risk groups. Furthermore, our analysis shows that PH patients are frequently diagnosed with other cardiovascular conditions, hypertension (58.1%) and atrial fibrillation (31.2%) being most common^1, 2, 3, 30^. Overlapping metabolic conditions such as diabetes, hypercholesterolemia, and atherosclerotic heart disease were prevalent, supporting the presence of cardiometabolic multimorbidity in PH patients. While these comorbidities are associated with worse outcomes, their mechanistic role in PH progression remains unclear and warrants further investigation. Although we did not directly measure systemic inflammation or vascular resistance, the observed comorbidity patterns, including high prevalence of metabolic conditions such as diabetes and hypercholesterolemia, suggest an underlying metabolic dysfunction that may contribute to disease progression in PH^29, 30, 31^. Supporting this is the genetic correlation between PH and several other cardiometabolic conditions, most notably heart failure, hypertension, atrial fibrillation, hypercholesterolemia, and diabetes. To test whether these comorbidities were predictive of PH diagnosis, a 12-features support vector machine model was trained on our matched case–control cohort and achieved a prediction AUC of 0.88 (Supplementary Figure S4). The most important features in the model were left heart failure, restrictive airway disease, valvular disease, atrial fibrillation, pulmonary embolism, and COPD (Supplementary Figure S5). This reinforces the prognostic importance of comorbidities in PH cases.

Process mining revealed critical time-based patterns in the progression of PH. The analysis shows that PH development and progression are often preceded by cardiovascular conditions such as hypertension and atrial fibrillation, alongside metabolic disorders like diabetes and hypercholesterolemia. These conditions frequently appear in diagnostic sequences before PH, suggesting potential shared risk factors or diagnostic complexity rather than direct causality ^32^. The quick decline in patient health after PH diagnosis highlights the critical need for prompt intervention and specialized treatment approaches to prevent negative health results ^33^. The sequence ordering of cardiometabolic comorbidities was associated with the speed of disease progression for the case of PH, COPD, and HF. For example, sequences beginning with diabetes followed by hypertension consistently led to earlier PH onset than the reverse order, highlighting a potential window for early screening in diabetic patients. This temporal sensitivity was mirrored in the COPD and HF cohorts, suggesting a generalizable pattern across cardiopulmonary conditions. Furthermore, the pathway *hypertension* → *atherosclerotic heart disease* → PH marked one of the fastest progression routes, with a median time to PH of 4 years and 11 months, compared to 5 years and 11 months when the order was reversed. This reinforces the role of early-onset hypertension as a key driver of accelerated cardiopulmonary deterioration. These findings highlight that hypertension preceding coronary artery disease may be a particularly aggressive progression route and could serve as an early warning signal for clinicians monitoring multimorbid patients. These findings align with earlier reports linking metabolic syndrome components to PH pathogenesis ^34^, but our results provide a longitudinal lens, demonstrating that sequence, not just the presence of comorbidities, carries important clinical implications.

For PH, the pathway *diabetes → hypertension → hypercholesterolemia → PH* showed the highest mortality risk which demonstrates the joint effect of diabetes, hypertension, and hypercholesterolemia. In HF, the pathway *hypertension → atrial fibrillation → diabetes →* HF exhibited the highest mortality risk which points to atrial fibrillation and hypertension as significant risk factors, especially when diabetes is present ^35^. These mortality-associated pathways highlight recurring patterns of metabolic and cardiovascular comorbidities, but further work is needed to determine whether sequence or individual conditions are key contributors to risk. There may also be beneficial outcomes linked to the comorbidity sequence, such as reduced mortality in pathways involving valvular diseases (*hypertension → mitral valve disorder →* PH) and chronic ischemic heart disease (*atherosclerotic heart disease → hypercholesterolemia →* HF). Early treatment of valvular dysfunction combined with hypercholesterolemia reduces negative health outcomes for patients with complex medical conditions ^35, 36, 37^. The occurrence rates of hypertension and atrial fibrillation within PH pathways match the research results established by Simonneau G *et. al.,* in ^1^, highlighted that left heart disease is the defining cause of PH in Group 2. Results from HF pathways demonstrate that successful arrhythmia management improves HF outcomes, which matches current evidence about atrial fibrillation protective effects ^38^. This analysis not only confirms the influence of known comorbidities but also highlights the possible distinct patterns of disease progression between PH and HF, emphasizing the need for condition-specific management strategies.

### Limitations

This study relied on ICD-10 codes to identify PH cases, which limits the ability to differentiate between specific PH subgroups (e.g., Group 1 PAH, Group 2 PH, Group 3 PH). Given the nature of hospital coding and the clinical profile of the UK Biobank population, it is likely that most PH cases represent Group 2 PH, with potential inclusion of some Group 3 cases. This assumption aligns with previous evidence suggesting that in large-scale population-based cohorts, PH diagnoses based on ICD-10 codes typically reflect Group 2 and Group 3 etiologies, particularly when comorbidities such as hypertension, atrial fibrillation, and COPD are prevalent ^38^. Additionally, the sex distribution in our PH cohort does not show the strong female predominance typically observed in Group 1 PH, further supporting the likelihood that most cases represent Group 2 or Group 3 PH. However, without confirmatory haemodynamic data or subgroup-specific diagnostic markers, precise classification is not possible. Treatment effects and interventions over time were not accounted for due to the high level of missingness in these records, potentially influencing disease trajectories. Future studies should address these gaps by incorporating treatment data, exploring diverse populations, and validating findings through external cohorts. Moreover, we should examine how treatment interventions, lifestyle modifications, and genetic and biomarker data influence disease pathways. Additional testing of machine learning models for disease-classification can incorporate clinical and comorbid diagnostic data associated with PH to provide a deeper understanding of long-term survival and quality of life.

## CONCLUSIONS

This study provides epidemiological evidence that the sequential order of cardiometabolic comorbidities influences disease progression and mortality in patients with PH, HF, and COPD. Using process mining and genetic correlation analysis, we showed that both the sequence and presence of conditions like diabetes, hypertension, and hypercholesteremia shape disease trajectories. These findings support the value of longitudinal risk assessment and highlight opportunities for earlier detection and intervention in multimorbid populations.

## Data Availability

This study was conducted using the UK Biobank data set under approved access (Application Number: 85457). Ethical approval was obtained from the Northwest Multi-center Research Ethics Committee (REC reference: 11/NW/0382), and all participants provided informed consent at recruitment. The data cleaning, preprocessing, and process mining stages of the analysis were implemented in Python (v 3.9.12) and R (v 4.3.2). The complete set of experiments, including the full analysis codebase for data processing, comorbidity trajectory mining, and figure generation, is permanently archived and freely accessible at: https://doi.org/10.5281/zenodo.15747766.

## Nonstandard Abbreviations and Acronyms

COPD: Chronic Obstructive Pulmonary Disease
GWAS: Genome-Wide Association Studies
HF: Heart failure
HR: Hazard Ratio
ICD: International Classification of Diseases
ILD: Interstitial Lung disease
PH: Pulmonary Hypertension

## ACKNOWLEDGEMENTS

This research was carried out using the UK Biobank Resource under Application Number 85457. We gratefully acknowledge the researchers and collaborators who provided support and guidance during this study.

All authors contributed to the study conception and design. Material preparation, data extraction, coding, analysis, and development of supplementary material were performed by Mohsin Masood. Hong Pan, and Naailah Bhamji supported coding and supplementary material. Genetic correlation analysis was carried out by Hong Pan. The first draft of the manuscript was written by Mohsin Masood. Juan Delgado-SanMartin, Siying Jane Ong, and Xiaohe Zhang contributed to manuscript refinement and editing. All authors read and approved of the final manuscript. Dennis Wang, Roger Thompson, and Allan Lawrie provided critical input on the research concept and contributed to manuscript review and refinement as co-senior authors.

